# Neural processes linking joint hypermobility and anxiety: Key roles for the amygdala and insular cortex

**DOI:** 10.1101/2024.10.10.24315234

**Authors:** Christina N Kampoureli, Charlotte L Rae, Cassandra Gould Van Praag, Neil A Harrison, Sarah N Garfinkel, Hugo D Critchley, Jessica A Eccles

## Abstract

**Background and Aims:** Anxiety symptoms are elevated among people with joint hypermobility. The underlying neural mechanisms are attributed theoretically to effects of variant connective tissue on the precision of interoceptive representations contributing to emotions.

**Methods:** We used functional magnetic resonance neuroimaging (fMRI) to quantify regional brain responses to emotional stimuli (facial expressions) in patients with generalised anxiety disorder (N=30) and a non-anxious comparison group (N=33). All participants were assessed for joint laxity and were classified (using Brighton Criteria) for the presence and absence of Hypermobility Syndrome (HMS: now considered Hypermobility Spectrum Disorder).

**Results:** HMS participants showed attenuated neural reactivity to emotional faces in specific frontal (inferior frontal gyrus, pre-supplementary motor area), midline (anterior mid and posterior cingulate cortices), and parietal (precuneus and supramarginal gyrus) regions. Notably, interaction between HMS and anxiety was expressed in reactivity of left amygdala (a region implicated in threat processing) and mid insula (primary interoceptive cortex) where activity was amplified in HMS patients with generalised anxiety disorder. Severity of hypermobility in anxious, compared to non-anxious, individuals correlated with activity within anterior insula (implicated as the neural substrate linking anxious feelings to physiological state). Amygdala-precuneus functional connectivity was stronger in HMS, compared to non-HMS, participants.

**Conclusions:** The predisposition to anxiety in people with variant connective tissue reflects dynamic interactions between neural centres processing threat (amygdala) and representing bodily state (insular and parietal cortices). Correspondingly, interventions to regulate of amygdala reactivity while enhancing interoceptive precision may have therapeutic benefit for symptomatic hypermobile individuals.

## Introduction

Joint hypermobility is one visible manifestation of familial connective tissue variants that can impact organ function throughout the body. Joint hypermobility often results in troublesome joint pain and stiffness (1) yet remains under-recognized and poorly managed. The diagnosis of joint hypermobility syndrome (HMS), as defined by the Brighton criteria, requires joint hypermobility plus musculoskeletal or connective tissue symptoms (e.g. prolapse, easy bruising, dislocations) (2). This classification has now been superseded by Hypermobility Spectrum Disorder (3). Rates of anxiety are markedly higher (odds ratio of 4.39) among hypermobile individuals (4). There is an over-representation of individuals with hypermobility in patients with anxiety-related conditions and in presentations in which anxiety frequently co-occurs, including neurodevelopmental conditions such as autism and ADHD (5,6). Furthermore, individuals with hypermobility often experience symptoms of dysautonomia, such as postural tachycardia syndrome (7). One explanation, connecting joint hypermobility, autonomic dysfunction and anxiety, proposes that relative inelasticity of connective tissue within peripheral vasculature compromises vasoconstriction and reduces venous return during standing through venous pooling. Compensatory autonomic responses, including increased sympathetic activity increases physiological arousal including heart rate (8–11).

In addition, hypermobile individuals may also show differences in interoceptive attention and sensitivity (increased sensing of changes from within the body (12)), reflecting the experience of greater interoceptive surprise through less predictable (more imprecise) afferent visceral signals (13). Increased attention may amplify interoceptive prediction error signals that contribute to the feeling of anxiety. Correspondingly, within the brain, hypermobile individuals are reported to show heightened reactivity in response to affective stimulation, both in regions responsible for interoceptive representation (insular cortex) and for emotional processing (amygdala) (14), where structural differences are even reported in people with sub-clinical hypermobile features (15).

The aim of this study is to use functional neuroimaging to explore the neural basis for the relationship between joint hypermobility and clinical anxiety, building on this earlier work (14,15). We hypothesized firstly that, participants with anxiety would exhibit heightened insula and amygdala reactivity when processing social emotional stimuli (facial expressions), replicating prior findings (13,16–18). No previous functional imaging work has specifically addressed the link between hypermobility and clinical anxiety, we therefore additionally hypothesised that the reactivity of amygdala and insula in conjunction with engagement of other ‘body-related’ brain regions would vary according to the presence and absence of hypermobility and anxiety, thereby illuminating neural substrates underlying their interaction (14,15).

## Method

### Participants

Sixty-three participants were recruited to the study. Patients volunteered to participate in response to an advertisement from Sussex Partnership NHS Trust either after inclusion in a linked study or via electronic bulletin boards. Members of the non-clinical comparison group were recruited via electronic bulletin boards. Of the sixty-three participants, thirty (47.6%) participants (age; mean ± standard error of the mean (SEM) = 42.93 yrs ± 2.24 yrs, 18 female, 12 male) met threshold for generalised anxiety disorder and thirty-three (52.4%) participants (age; mean ± SEM = 37.42 yrs ± 2.28, 16 female, 17 male) were healthy controls. There were no statistically significant differences in age or gender between the two groups. Of the patients with generalized anxiety disorder, eighteen (60%) were classified as having joint hypermobility syndrome, and seven (21.2%) of the non-clinical comparison group met diagnostic threshold for joint hypermobility syndrome. See Table 1 for participant demographic details and clinical features.

**Table 1.** Demographic details and anxiety level (measured using the Beck Anxiety Inventory-BAI) of participants with generalized anxiety disorder (GAD) and the non-clinical comparison group. Group difference p-values refer to a two-tailed t-test (age) or chi-squared test (sex).

| Group | GAD:<br>HMS | GAD:<br>non-HMS | Comparison:<br>HMS | Comparison:<br>non-HMS | Group<br>difference |
| --- | --- | --- | --- | --- | --- |
| Number | 18 | 12 | 7 | 26 |  |
|  | 30 |  | 33 |  |  |
| Age<br>(mean, SEM) | 43 ± 2 | | 37 ± 2 | | $t(61) = -1.72$,<br>$p = .091$ |
| Sex | 18 female; 12 male | | 16 female; 17 male | | $\chi^2(1) = .84$,<br>$p = .360$ |
| BAI<br>(mean, SEM) | 22.9(1.78) | | 5.48(.70) | | $t(61) = -9.11$<br>$p < .001$ |

Inclusion criteria for patients included DSM-IV diagnosis of generalized anxiety disorder. Participants in the non-clinical comparison group were required to be free from any history of psychiatric disorder. General exclusion criteria included MRI incompatibility, presence of neurological illness, and presence of diagnosed psychiatric illness other than anxiety or co-morbid depression in patients. Written informed consent was obtained from all participants. The authors assert that all procedures contributing to this work comply with the ethical standards of the relevant national and institutional committees on human experimentation and with the Helsinki Declaration of 1975, as revised in 2008. All procedures involving human participants including patients were approved by the National Research Ethics Service – South East Coast (Brighton and Sussex; REC Reference 12/LO/1942; IRAS registration number 115219).

All participants underwent assessment for generalized anxiety disorder and hypermobility. DSM-IV diagnosis of generalized anxiety disorder was confirmed or refuted using the MINI (The Mini – International Neuropsychiatric Interview) (19). The presence or absence of generalised joint laxity was established through physical examination of joints using the Beighton Scale (20) where a cut off of 4 out of 9 was used in line with the UK literature e.g. (21). All participants were assessed by the same clinician (J.A.E.). The presence of hypermobility syndrome (HMS) was confirmed or excluded using Brighton Criteria(2).

Anxiety level was measured using the Beck Anxiety Inventory (BAI)(22).There was significant group difference in anxiety levels (BAI; mean ± SEM: anxious group 22.9 ± 1.78 vs non-anxious group 5.48 ± 0.70; ***t*** (61) = 9.11, ***p*** < .001). See Table 1 for anxiety levels measured using the BAI across groups. There was no significant difference in anxiety level between those with hypermobility syndrome and those without, either in the anxious (mean ± SEM: anxious group:hms 23.28 ± 2.37 vs anxious group: non-hms 22.42 ± 2.82; ***t*** (28) = - .23, ***p*** = .818) or non-anxious group (non-anxious group:hms 5.38 ± .74 vs non-anxious group:non-hms 5.86 ± 1.92; ***t*** (31) = -.27, ***p*** = .786). All clinical and demographic data was analysed with IBM SPSS Statistics v. 29(23).

### Emotional faces task

An emotional faces task was modified from Umeda and colleagues (24), wherein five classes of images of emotional faces from the Karolinska Directed Emotional Faces set (KDEF) (classes: angry, afraid, disgusted, happy, and neutral) (25), were presented in a randomised order. Null events were pseudo-randomised and presented as fixation cross. These were also included to facilitate the identification of haemodynamic responses to stochastically ordered stimuli. There were fifteen trials of each emotion category, and twenty-one null events, each lasting four seconds. During each face presentation, participants were asked to make an incidental judgement of whether they could see teeth or not (index or middle finger button press with right hand), to ensure attention to the stimuli.

### MRI acquisition

Functional MRI data were acquired on a Siemens Avanto 1.5 Tesla with a 32-channel head coil (T_2_*-weighted echo planar images, repetition time = 2520 ms, echo time = 43 ms, 34 interleaved slices 3-mm thick with 0.6 mm interslice gap, in-plane resolution 3 × 3 mm). A T_1_ structural was acquired for registration (repetition time = 2730 ms, echo time = 3.57 ms, 1×1×1 mm resolution).

### fMRI preprocessing

Functional MRI data were pre-processed and analysed using Statistical Parametric Mapping (SPM12)(26) running in MATLAB(27). Pre-processing was performed using default options, including realignment to the mean image, slice-time correction to the 6^th^ slice, co-registration to the T1 structural image, normalization to MNI space, as well as smoothing at 8 mm Gaussian smoothing kernel. To account for head motion, framewise displacement (FD) values were calculated from the six motion parameters (rp_.txt) generated in SPM during realignment using the FD_conn method in the CONN toolbox (version 22.a)(28). Participants with excessive motion (FD > 0.5 mm) were flagged for closer inspection. Two participants exceeded this threshold (with FD values of 0.63 mm and 0.60 mm, respectively) but were retained in the analysis to preserve the representativeness and generalizability of the findings within the clinical population.

### Statistical analysis

#### First-level general linear model

Task events were modelled in a general linear model, with five regressors representing the onset and duration of presentation of angry, afraid, disgusted, neutral and happy faces respectively. To account for head motion, six nuisance regressors modelled head movement using the motion parameters calculated during realignment. Single-regressor T-contrasts were generated for viewing (i) angry, (ii) afraid, (iii) disgusted, (iv) neutral and (v) happy faces by assigning a contrast weight of 1 to each of the five experimental conditions, with the intertrial interval fixation cross representing an implicit baseline. These T-contrasts were entered to a full factorial second-level analysis.

#### Second-level general linear model

A second-level full factorial model contained HMS (non-HMS, HMS) and anxiety (non-anxious, anxious) as an independent (between-subjects) factor, and facial expression (angry, afraid, disgusted, neutral and happy) as a within subject factor. In addition, two covariates were entered for (i) gender (male, female) and (ii) age.

F-contrasts were generated testing for: all effects, main effect of HMS, main effect of anxiety, main effect of task, and interactions between the factors. Individual group effects for viewing faces (compared to implicit baseline) were examined using T-contrasts: HMS> non-HMS; HMS (anxious>non-anxious); non-HMS>HMS; and anxious (HMS>non-HMS). A series of two further second-level models included Beighton score and anxiety level (BAI score) as additional covariates. In the Beighton second-level model, the Beighton score was used as a covariate so that the main effect of hypermobility symptoms could be modelled along with the interaction of hypermobility symptoms with the anxiety factor (i.e. presence of GAD or not). In the anxiety second-level model, anxiety level (BAI score) was used as a covariate, so that the main effect of anxiety level could be modelled along with the interaction of anxiety level (BAI score) with the factor of HMS. All covariates were mean centred around zero.

Statistical images were thresholded at a cluster-forming threshold of *P* < 0.001 for family-wise error rate (FWEc) correction for multiple comparisons at *P* < 0.05 (29). Significant clusters were localized according to the Anatomy toolbox (v 3.0)(30).

### Psychophysiological interactions

We performed a series of psychophysiological interaction analyses to investigate how brain activity in response to emotional faces, within regions identified in the above univariate analyses, changed in their functional connectivity to other regions of the brain as a function of hypermobility and anxiety status. On the basis of the univariate fMRI results, we identified 3 regions from which to seed these functional connectivity analyses: (i) left amygdala (centred on x -32, y 0, z -16); (ii) right mid insula (centred on x 34, y -2, z -6); and (iii) left inferior frontal gyrus (x -46, y 34, z 2). Firstly, we extracted the first eigenvariate (weighted mean of BOLD time series), for each region by thresholding three contrasts at *P* < 1 for each participant: (i) the interaction between anxiety and HMS (for left amygdala ROI); (ii) the main effect of HMS for (right mid insula ROI); and (iii) the main effect of HMS (for left inferior frontal gyrus ROI). Then, an F-contrast was computed for each subject, representing all effects (angry, afraid, disgusted, neutral and happy: “eye:5”). In the three contrasts given above, we then extracted a 10 mm sphere of voxels for each ROI, adjusting for the F-contrast of all effects.

Next, the psychophysiological interaction term was calculated according to the main effect of task (contrast weights: 1 for angry, 1 for afraid, 1 for disgusted, 1 for neutral and 1 for happy) and the BOLD time series for each ROI. These psychophysiological interaction terms were each entered into a first-level model for each participant, alongside a regressor representing the BOLD activity of the ROI (PPI.Y) and the main effect of task (PPI.P). Single regressor T-contrasts were generated for the psychophysiological interaction term using a single contrast weight to investigate positive changes in the regression slope of voxels elsewhere in the brain relative to the seed ROI during task events relative to baseline.

The first-level T-contrasts were then entered to a series of second-level models that examined the psychophysiological interaction between the seed ROI and voxels across the brain using a full factorial second-level analysis, with HMS (non-HMS, HMS) and anxiety (non-anxious, anxious) as an independent (between-participant) factor, and the first-level T-contrasts representing functional connectivity when viewing faces as a repeated measures (within-participant) factor.

In these second-level models, as with the univariate functional MRI analysis, age and gender were entered as covariates. Contrasts were thresholded at a cluster-forming threshold of *P* < 0.001 for family-wise FWEc at *P* < 0.05. Significant clusters were localized with reference to the SPM Anatomy toolbox (v 2.2b)(30).

### Transparency Declaration

We affirm that the manuscript is an honest, accurate, and transparent account of the study being reported and that no important aspects of the study have been omitted.

### Analytic Code Availability

The analytic code (SPM batches) that was used for the neuroimaging analysis for this study is available at OSF: DOI 10.17605/OSF.IO/TCEMX.

### Research Material and Data Availability

The MRI acquisition sequence information, demographic and clinical data, and participant mean FD values are available at OSF: DOI 10.17605/OSF.IO/TCEMX. The neuroimaging data that support the findings of this study (unthresholded statistic images for every contrast reported) are openly available at https://identifiers.org/neurovault.collection:16863/, reference number 16863(31).

## Results

### Univariate functional MRI

#### Main effects

As anticipated, there was a significant main effect of anxiety within the left amygdala and right mid insula, with post hoc T-contrasts revealing greater activity in these regions in anxious versus non-anxious participants (Supplementary Material, Table 4 & 5).

We also observed significant main effects of HMS in left inferior frontal gyrus, precuneus and pre-SMA, right mid insula, right posterior and left anterior mid cingulate gyrus and the left supramarginal gyrus (Supplementary Material, Table 1). *Post hoc* T-contrasts revealed that non-HMS participants showed greater activity in left inferior frontal gyrus, precuneus, left pre-SMA, right posterior and left anterior mid cingulate gyrus, the left supramarginal gyrus when viewing emotional faces compared to HMS participants (Figure 1A). *Post hoc* T-contrasts for the right mid insula were not statistically significant.

**Figure 1.**
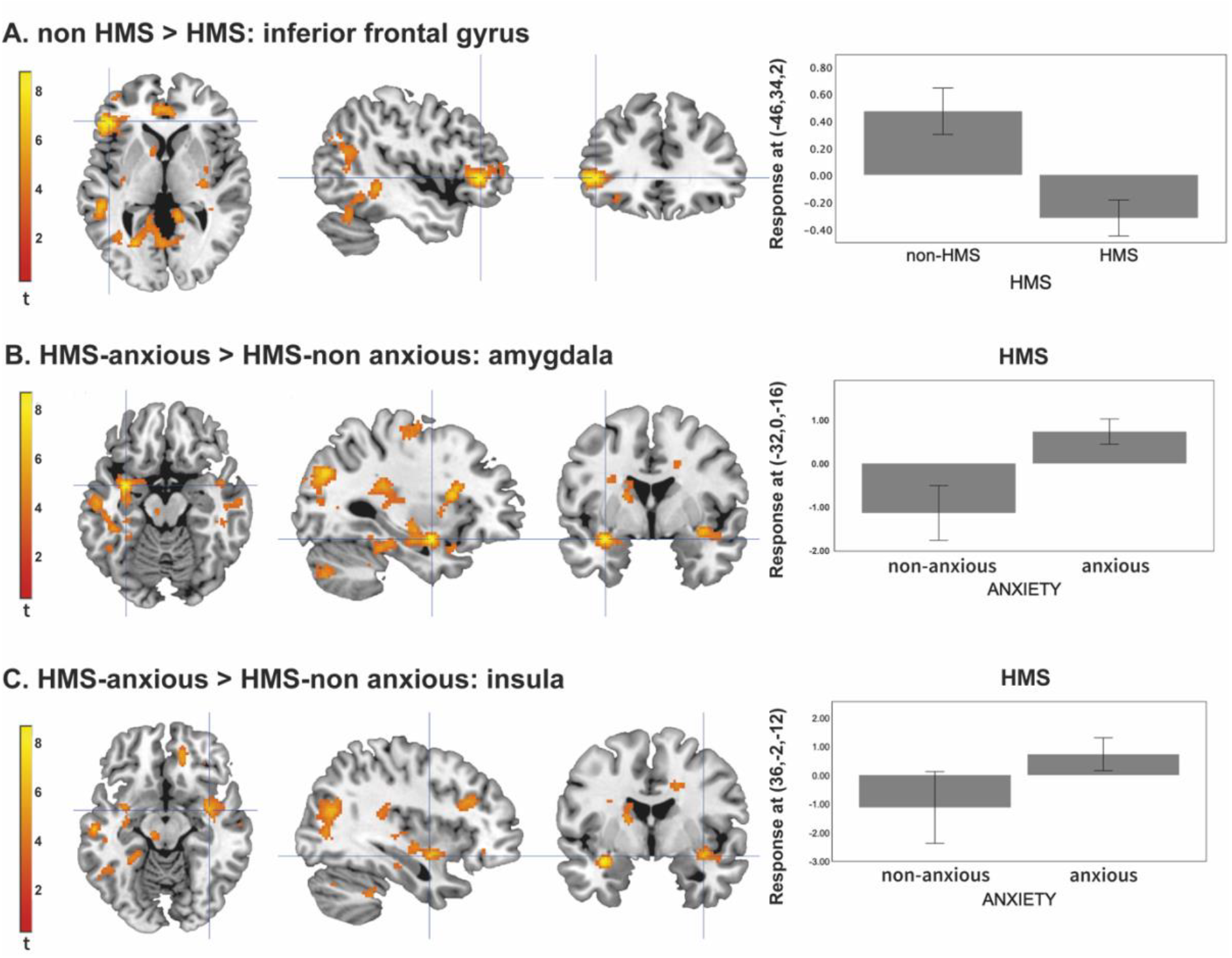
Activity during viewing emotional faces. A. Activation in the inferior frontal gyrus is greater in non-HMS participants compared to the HMS. B. Activation in the amygdala is greater in the HMS anxious participants compared to the HMS non-anxious participants. C. Activation in the mid-insula is greater in the HMS anxious participants compared to the HMS non-anxious participants. Unthresholded statistic images are openly available in Neurovault, at https://identifiers.org/neurovault.collection:16863/. HMS refers to the diagnosis of Joint Hypermobility Syndrome (HMS).

We also observed the known main effect of viewing emotional faces, associated with activation of large areas of the occipital lobe, right middle and left superior frontal gyrus (Supplementary Material, Table 3).

#### Interaction: Hypermobility x Anxiety

Furthermore, there was a significant interaction between HMS and anxiety in left amygdala, left hippocampus, right paracingulate gyrus and right mid insula (Supplementary Material, Table 6). *Post hoc* T-contrasts revealed: 1) that in the HMS group, there was greater activation in the left amygdala and the right paracingulate gyrus in anxious compared to non-anxious participants (Figure 1B); findings for the left hippocampus were not significant. 2) In participants with anxiety, there was greater activation in the right paracingulate gyrus and right mid-insula in HMS compared to the non-HMS (Figure 1C).

#### Interaction: Beighton score x anxiety

The interaction between number of hypermobile joints (Beighton score) and anxiety status, i.e. the interaction testing for regions in which activation was more positively correlated with Beighton score for anxious compared to non-anxious participants, showed activation in the left anterior insula (Figure 2A).

**Figure 2.**
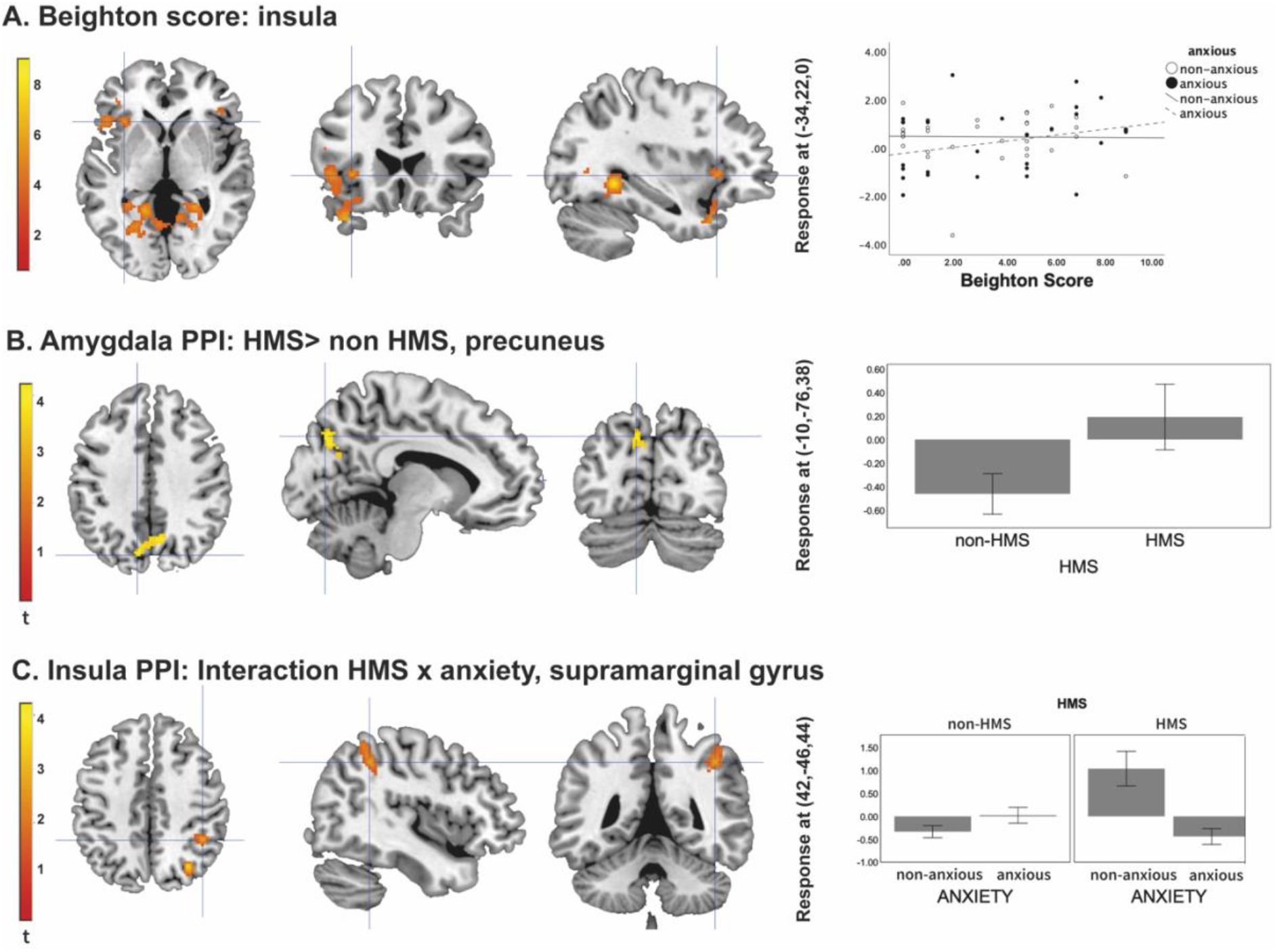
A. Activation in anterior insula: interaction between Beighton score and anxiety status. B. Changes in functional connectivity between the amygdala and the precuneus in HMS participants compared to non-HMS. C. Changes in functional connectivity between the insula and supramarginal gyrus and the interaction between HMS and anxiety. Unthresholded statistic images are openly available in Neurovault, at https://identifiers.org/neurovault.collection:16863/. HMS refers to the diagnosis of Joint Hypermobility Syndrome (HMS).

#### Interaction: Beck anxiety score x Hypermobility

The interaction between anxiety severity (Beck Anxiety Inventory score) and HMS, i.e., the interaction testing for regions in which activation was more positively correlated with anxiety score for HMS compared to non-HMS participants, showed activation in the left putamen (Supplementary Material, Table 10).

### Psychophysiological Interactions

Three second-level models examined changes in functional connectivity with (i) the left amygdala; (ii) the right mid insula and (iii) the left inferior frontal gyrus, depending on the psychological context of viewing emotional faces. In the left amygdala psychophysiological interaction, there was no significant effect of HMS (F-contrast; HMS, non-HMS). However, the *post-hoc* T-contrast testing for HMS versus non-HMS (T-contrast; HMS> non-HMS [ -1 - 1 1 1]) revealed that HMS participants showed greater functional connectivity when viewing faces between the left amygdala and the left precuneus (Figure 2B).

In the right mid insula psychophysiological interaction, there was a significant interaction effect between HMS and anxiety severity in the right supramarginal gyrus (Figure 2C), and the right occipital cortex. However, the *post hoc* T-contrasts to explore the effect did not remain significant after correction for multiple comparisons. The left inferior frontal gyrus psychophysiological interaction did not produce statistically significant results.

## Discussion

Here, using functional neuroimaging during the incidental processing of emotional faces, we identify putative neural substrates underpinning the association between hypermobility and anxiety. Hypermobile participants showed reduced activation in discrete areas of association cortex, notably prefrontal and parietal regions. Anxious participants, in line with several previous imaging studies, showed amplified reactivity of the amygdala and insula during a socio-emotional challenge. For the first time, we were able to investigate the neural interaction between hypermobility and anxiety. We first confirmed that anxious HMS participants showed greater activity in the amygdala and insula than non-anxious HMS participants. Furthermore, the degree of hypermobility (as measured with Beighton score) was more strongly correlated with insula activity in anxious HMS participants than non-anxious HMS participants. Finally, there was a general effect of hypermobility (regardless of anxiety) on functional connectivity between the amygdala and precuneus. Collectively, these findings identify a network of amygdala, insula, and association cortices that link anxiety and hypermobility. Greater connectivity and activity within this network may underpin the increased prevalence of anxiety in individuals with hypermobility syndrome.

### Interoceptive pathways in the brain

The brain continuously receives sensory information from the visceral organs and peripheral tissues via ascending nerves that enter the brainstem (32). From here, interoceptive signals relaying the state of the body are conveyed to the thalamus, and ultimately posterior insula cortex. Here, viscerotopic representations of these afferent signals are believed to support the cognitive perception of bodily feelings (e.g. heart rate, respiration, gastric sensations). This viscerotopic information is then re-represented, and integrated, more anteriorly in the insula lobe, underpinning our experience of broader affective states (33,34).

Putatively, hypermobility may render individuals more prone to anxious affective experiences via heightened signalling of interoceptive signals relaying dysautonomic states. Due to changes in the connective tissue of the vasculature, hypermobile people may experience abnormal peripheral vasoconstriction(35). Specifically, reduced venous return during standing due to venous pooling may be responsible for an increased sympathetic state, and autonomic hyperactivity (8,9). The insular cortex is an important central substrate for receiving this autonomic hyperactivity information.

It is therefore particularly intriguing that the insula was not only identified as overactive in our anxious sample, but more specifically, as more active in anxious versus non-anxious HMS participants. We also saw that the degree of hypermobility (Beighton score) was more strongly correlated with insula activity in anxious than non-anxious HMS participants. This identifies the insula as a nexus of affective experience in anxious hypermobile people. This finding extends similar observations reported in other affective conditions (36)(e.g. increased emotional reactivity (hyperactivity of salience-processing regions) in bipolar disorder) and previous findings from our group that have examined the functional activity in non-anxious hypermobile participants (14). Within this context, interoception, i.e. the dynamic signalling, neural and perceptual representation of internal physiological states of the body, is a likely unifying factor. Additionally, patients with bipolar disorder have demonstrated abnormal insular functional connectivity, possibly modulated by inflammatory markers (37). Similar mechanisms may underpin the heightened insula activity observed in anxious hypermobile participants, potentially mediated by dysautonomic states.

### Amygdala interactions with hypermobility

In addition to reactivity differences in the insular cortex, we also observed interesting findings in the amygdala. Affective tasks performed during fMRI reliably engage the amygdala. This activation is typically amplified in anxious participants (38) and is also enhanced following interoceptive stressors such as immune challenges (39). It is thus noteworthy that the amygdala was identified as a functional neural centre for interaction between anxiety and hypermobility, wherein the amygdala showed even greater activity in anxious HMS participants than the non-anxious HMS group. The amygdala is a critical region supporting the detection and perception of threat through associative integration of external and interoceptive information (40), which may underpin its role in affective experience(41). The greater reactivity of the amygdala in anxious hypermobile individuals may also reflect previously identified differences of amygdalar structure and function in hypermobile people (14,15). In parallel, the amygdala response in anxious HMS participants may reflect the dynamic contributions (e.g. to behavioural / autonomic response and subjective feelings) of amygdala and the insula within a wider affective network; although our functional connectivity analyses did not identify a dependent association between these two regions.

Functional connectivity analyses did however identify stronger functional coupling between amygdala and precuneus in hypermobile participants (regardless of anxiety). This may suggest that hypermobile individuals have a tendency towards hyper-connectivity of affective regions, which may have consequences for onward processing of information.

### Affective tasks as a probe for functional anatomy

We selected an emotional face processing task as a vehicle for probing affective responses in participants with generalised anxiety disorder. A wide literature has previously taken such an approach to confirm the involvement of regions such as the amygdala and insula in a variety of anxiety conditions(42) as well as experimental interoceptive challenges (39). Here, we were able to extend this literature to understand the impact of hypermobility on these processes.

In our analyses, we collapsed across the five stimuli types to ask the fundamental question of how viewing social affective stimuli can provoke a neural response. In future work, it would be interesting to tease apart further the nuances of different affective cues on the hypermobile brain. For example, do responses to anger differ to fear?

### Limitations and future directions

Despite our implementation of a well-established paradigm to invokes reliable activations in affective brain regions, we acknowledge that fMRI tasks constrain one’s investigative potential to the circumscribed set of regions that the task recruits. Other approaches, including resting-state fMRI provide complementary information that can be leveraged to examine brain-wise network interactions. We did not acquire such data within this study, yet this remains an important avenue for future work to understand better the neural characteristics of hypermobility.

We recruited a community sample that enabled us to screen participants and place them into one of four categories according to anxiety and hypermobility status. However, since hypermobility is a risk factor for anxiety, with an odds ratio of 4.39 for suffering from anxiety if hypermobile (4), recruiting large numbers of hypermobile individuals who are not anxious (and anxious individuals who are not hypermobile) was a challenge. This means our sample sizes for the non-anxious HMS and anxious non-HMS groups were smaller than the other two groups.

Future work should capitalise on advances in biofeedback interventions (such as ADIE(43) and ADAPT(44)) to target anxiety in hypermobile individuals, perhaps using neurofeedback to specifically down-regulate insular and amygdala reactivity. As in the broader mental health space, it is becoming increasingly clear that a ‘one size fits all,’ approach does not work for many groups who experiencing anxiety: The present study will potentially inform personalised treatment approaches (45) for a group of individuals who have previously perhaps been dismissed or overlooked(46).

## Conclusions

We identified a putative neural network that underpins the link between joint hypermobility and anxiety. The insula and amygdala are critical hubs that show a greater response to affective stimuli in anxious hypermobile individuals. Our findings highlight these regions as potential targets for therapeutic interventions such as biofeedback that an prove crucial for an often under-recognised, overlooked by significant proportion of anxiety patients.

## Data Availability

Analytic Code Availability:
The analytic code (SPM batches) that was used for the neuroimaging analysis for this study is available at OSF: DOI 10.17605/OSF.IO/TCEMX.
Research Material and Data Availability:
The MRI acquisition sequence information, demographic and clinical data, and participant mean FD values are available at OSF: DOI 10.17605/OSF.IO/TCEMX. The neuroimaging data that support the findings of this study (unthresholded statistic images for every contrast reported) are openly available at https://identifiers.org/neurovault.collection:16863/, reference number 16863(31).

https://identifiers.org/neurovault.collection:16863/

## CRediT Author contributions

CNK: Methodology, Formal Analysis, Visualization, Writing – original draft, Writing – review & editing; CLR: Methodology, Formal Analysis, Supervision, Writing – original draft, Writing – review & editing; CGvP: Methodology, Writing – review & editing; NAH: Writing – review & editing; SNG: Writing – review & editing; HDC: Conceptualization, Supervision, Writing – review & editing; JAE: Conceptualization, Project Administration, Methodology, Investigation, Data Curation, Formal Analysis, Supervision, Writing – original draft, Writing – review & editing.

## Acknowledgements

We would like to thank the radiography team and administrative staff at the Clinical Imaging Sciences Centre (CISC) for all their support throughout this project.

