## Supplementary Material for "Neural processes linking joint hypermobility and anxiety: Key roles for the amygdala and insular cortex"

| **Table 1. Main effect of HMS (F-contrasts) (FEWc: 106, c:32)** | | | | | | | | |
| --- | --- | --- | --- | --- | --- | --- | --- | --- |
| equivK | F | equivZ | x | y | | z | Label | Hemisphere |
| 393 | 77.72 | Inf | -46 | 34 | 2 | | inferior frontal gyrus pars triangularis | L |
| 922 | 53.85 | 6.93 | 0 | -52 | 38 | | precuneus cortex |  |
| 485 | 42.60 | 6.19 | -2 | 0 | 56 | | pre-SMA | L |
| 591 | 58.40 | 7.20 | 34 | -2 | -6 | | Mid Insular cortex | R |
| 228 | 43.61 | 6.26 | 2 | -50 | 16 | | cingulate gyrus posterior division | R |
| 167 | 36.45 | 5.74 | -2 | 44 | 4 | | cingulate gyrus anterior division | L |
| 177 | 48.33 | 6.58 | -56 | -32 | 40 | | supramarginal gyrus, anterior division | L |
| 616 | 67.00 | 7.67 | 22 | -40 | -16 | | temporal occipital fusiform cortex | R |
| 1767 | 61.02 | 7.35 | -26 | 12 | 40 | | cerebral white matter | L |
| 370 | 57.11 | 7.12 | -34 | -2 | -14 | | WM Inferior occipito-frontal fascicle | L |
| 350 | 55.03 | 7.00 | -34 | -26 | 26 | | parietal operculum cortex | L |
| 2520 | 53.51 | 6.91 | -50 | -38 | -6 | | middle temporal gyrus posterior division | L |
| 106 | 53.19 | 6.89 | 30 | -72 | -6 | | occipital fusiform gyrus | R |
| 254 | 52.72 | 6.86 | 10 | 18 | 50 | | superior frontal gyrus | R |
| 183 | 46.51 | 6.46 | -34 | -28 | -14 | | left hippocampus | L |
| 364 | 43.53 | 6.26 | -20 | -14 | 74 | | precentral gyrus | L |
| 132 | 40.45 | 6.04 | -32 | -54 | -34 | | cerebellum left VI | L |
| 304 | 40.11 | 6.01 | -20 | 40 | 20 | | frontal pole | L |
| 249 | 40.08 | 6.01 | 26 | -46 | 48 | | superior parietal lobule | R |
| 295 | 39.91 | 6.00 | -42 | -2 | -26 | | temporal pole | L |
| 142 | 38.47 | 5.89 | -60 | -18 | 4 | | planum temporale | L |
| 275 | 38.28 | 5.88 | 14 | 22 | 18 | | right caudate | R |
| 246 | 37.95 | 5.85 | 0 | -70 | -18 | | cerebellum vermis vi |  |
| 162 | 31.52 | 5.34 | -22 | 44 | -8 | | frontal pole | L |
| 186 | 31.45 | 5.34 | -40 | -76 | 14 | | lateral occipital cortex inferior division | L |
| 175 | 31.16 | 5.31 | -50 | -54 | 20 | | angular gyrus | L |
| 147 | 31.08 | 5.31 | 52 | 4 | 8 | | precentral gyrus | R |
| 245 | 30.46 | 5.25 | 24 | -2 | 30 | | cerebral white matter | R |
| 167 | 29.02 | 5.13 | -14 | -82 | 44 | | lateral occipital cortex superior division | L |
| 140 | 27.99 | 5.03 | 0 | 6 | -8 | | left caudate |  |
| 159 | 25.66 | 4.82 | 48 | 8 | 22 | | precentral gyrus | R |

| **Table 2. Main effect of HMS (Post- hoc T-contrasts) controls>hms (FEWc:166, c:17)** | | | | | | | |
| --- | --- | --- | --- | --- | --- | --- | --- |
| equivK | T | equivZ | x | y | z | Label | Hemisphere |
| 642 | 8.82 | Inf | -46 | 34 | 2 | Inferior frontal gyrus, pars triangularis | L |
| 1113 | 7.34 | 7.02 | 0 | -52 | 38 | precuneus cortex |  |
| 565 | 6.53 | 6.30 | -2 | 0 | 56 | pre-SMA | L |
| 255 | 6.60 | 6.37 | 2 | -50 | 16 | cingulate gyrus posterior division | R |
| 192 | 6.04 | 5.86 | -2 | 44 | 4 | cingulate gyrus anterior division | L |
| 209 | 6.95 | 6.68 | -56 | -32 | 40 | supramarginal gyrus, anterior division | L |
| 1477 | 8.19 | 7.75 | 22 | -40 | -16 | temporal occipital fusiform cortex | R |
| 666 | 7.56 | 7.22 | -34 | -2 | -14 | temporal fusiform cortex posterior division | L |
| 3284 | 7.32 | 7.00 | -50 | -38 | -6 | middle temporal gyrus, posterior division | L |
| 166 | 6.97 | 6.70 | 42 | -44 | 10 | middle temporal gyrus, temporooccipital | R |
| 465 | 6.60 | 6.37 | -20 | -14 | 74 | precentral gyrus | L |
| 311 | 6.33 | 6.12 | 26 | -46 | 48 | superior parietal lobule | R |
| 232 | 5.61 | 5.46 | -40 | -76 | 14 | lateral occipital cortex, inferior division | L |
| 227 | 5.58 | 5.44 | -50 | -54 | 20 | angular gyrus | L |
| 170 | 5.57 | 5.43 | 52 | 4 | 8 | precentral gyrus | R |
| 208 | 5.39 | 5.26 | -14 | -82 | 44 | lateral occipital cortex, superior division | L |
| 171 | 5.29 | 5.17 | 0 | 6 | -8 | left caudate |  |

| **Table 3. Main effect of Task (F-contrasts) (FEWc: 108, c:13)** | | | | | | | |
| --- | --- | --- | --- | --- | --- | --- | --- |
| equivK | F | equivZ | x | y | z | Label | Hemisphere |
| 5569 | 211.48 | Inf | -40 | -76 | 32 | lateral occipital cortex superior division | L |
| 407 | 95.86 | Inf | 28 | 26 | 40 | middle frontal gyrus | R |
| 271 | 82.73 | Inf | -20 | 26 | 42 | superior frontal gyrus | L |
| 75061 | 715.62 | Inf | 10 | -82 | -4 | lingual gyrus v2 | R |
| 210 | 72.69 | Inf | -62 | -12 | -14 | middle temporal gyrus posterior division | L |
| 238 | 65.52 | 7.59 | -62 | -56 | -4 | middle temporal gyrus, temporooccipital | L |
| 719 | 58.11 | 7.18 | -8 | 42 | -12 | frontal medial cortex | L |
| 408 | 52.01 | 6.81 | -40 | -10 | -26 | temporal fusiform cortex, posterior division | L |
| 214 | 49.44 | 6.65 | -20 | -26 | 36 |  | L |
| 118 | 36.99 | 5.78 | -24 | 8 | 26 | left caudate | L |
| 108 | 29.26 | 5.15 | 30 | -42 | -6 | lingual gyrus | R |
| 120 | 28.25 | 5.06 | -28 | -42 | -8 | lingual gyrus | L |
| 155 | 28.01 | 5.04 | 20 | -80 | -34 | cerebellum right crus ii | R |

| **Table 4. Main effect of Anxiety (F-contrasts) (FWEc: 103, c: 30)** | | | | | | | |
| --- | --- | --- | --- | --- | --- | --- | --- |
| equivK | F | equivZ | x | y | z | Label | Hemisphere |
| 289 | 50.85 | 6.74 | -32 | -2 | -16 | left amygdala | L |
| 244 | 49.09 | 6.63 | 38 | -2 | -10 | Mid insular cortex | R |
| 817 | 64.09 | 7.52 | -22 | -12 | 38 |  | L |
| 145 | 55.73 | 7.04 | 16 | 30 | 4 |  | R |
| 158 | 52.85 | 6.87 | -50 | -12 | 6 | heschl’s gyrus | L |
| 346 | 49.18 | 6.64 | -26 | -64 | -40 | cerebellum left crus i | L |
| 1284 | 47.10 | 6.50 | -8 | -28 | 58 | precentral gyrus | L |
| 157 | 44.99 | 6.36 | -34 | 8 | 30 | middle frontal gyrus | L |
| 133 | 44.07 | 6.30 | -46 | 32 | 4 | inferior frontal gyrus, pars triangularis | L |
| 337 | 42.72 | 6.20 | 16 | 36 | -10 | frontal pole | R |
| 134 | 42.29 | 6.17 | 30 | -86 | -2 | lateral occipital cortex, inferior division | R |
| 313 | 41.80 | 6.14 | -32 | 16 | 14 | frontal operculum cortex | L |
| 248 | 41.58 | 6.12 | -2 | -48 | -14 | cerebellum left i-iv | L |
| 295 | 41.54 | 6.12 | 14 | 8 | 58 | superior frontal gyrus | R |
| 184 | 39.80 | 5.99 | 32 | 6 | 32 | precentral gyrus | R2 |
| 725 | 39.51 | 5.97 | -24 | -66 | 4 | intracalcarine cortex | L |
| 114 | 36.98 | 5.78 | 16 | -38 | -22 | cerebellum right i-iv | R |
| 472 | 35.82 | 5.69 | 32 | -88 | 16 | lateral occipital cortex, superior division | R |
| 195 | 35.74 | 5.68 | -10 | 50 | 10 | paracingulate gyrus | L |
| 188 | 35.34 | 5.65 | 4 | 18 | 52 | superior frontal gyrus | R |
| 103 | 34.55 | 5.59 | -24 | -32 | 4 | left thalamus | L |
| 264 | 34.08 | 5.55 | 12 | -56 | 50 | precuneus cortex | R |
| 312 | 33.45 | 5.50 | -50 | -58 | -30 | cerebellum left crus i | L |
| 183 | 33.30 | 5.49 | 22 | -72 | -36 | cerebellum left crus i | L |
| 173 | 31.62 | 5.35 | 54 | 8 | 6 | precentral gyrus | R |
| 273 | 30.34 | 5.24 | 40 | 22 | 26 | middle frontal gyrus | R |
| 165 | 29.92 | 5.21 | 48 | -20 | 38 | postcentral gyrus | R |
| 152 | 29.41 | 5.16 | 14 | -28 | -12 | brain stem | R |
| 144 | 29 | 5.16 | -66 | -38 | 0 | middle temporal gyrus, posterior division | L |
| 143 | 26.93 | 4.94 | -20 | -80 | 42 | lateral occipital cortex, superior division | L |

| **Table 5. Main effect of Anxiety (Post- hoc T-contrasts) anx>non-anx (FEWc:123, c:19)** | | | | | | | |
| --- | --- | --- | --- | --- | --- | --- | --- |
| equivK | T | equivZ | x | y | z | Label | Hemisphere |
| 422 | 7.13 | 6.84 | -32 | -2 | -16 | left amygdala | L |
| 127 | 7.04 | 6.76 | 48 | -50 | -40 | cerebellum | R |
| 770 | 7.01 | 6.74 | -26 | -64 | -40 | cerebellum | L |
| 277 | 7.01 | 6.73 | 38 | -2 | -10 | Mid insular cortex | R |
| 2726 | 6.86 | 6.60 | -8 | -28 | 58 | precentral gyrus | L |
| 154 | 6.64 | 6.40 | -46 | 32 | 4 | inferior frontal gyrus | L |
| 401 | 6.54 | 6.31 | 16 | 36 | -10 | frontal pole | R |
| 356 | 6.46 | 6.25 | -32 | 16 | 14 | frontal operculum cortex | L |
| 296 | 6.45 | 6.23 | -2 | -48 | -14 | cerebellum | L |
| 142 | 6.20 | 6.00 | 42 | -44 | 8 | supramarginal gyrus | R |
| 147 | 6.08 | 5.90 | 16 | -38 | -22 | cerebellum | R |
| 554 | 5.98 | 5.81 | 32 | -88 | 16 | lateral occipital cortex superior division | R |
| 245 | 5.98 | 5.80 | -10 | 50 | 10 | paracingulate gyrus | L |
| 311 | 5.84 | 5.67 | 12 | -56 | 50 | precuneus cortex | R |
| 225 | 5.77 | 5.61 | 22 | -72 | -36 | cerebellum | L |
| 204 | 5.62 | 5.48 | 54 | 8.0 | 6 | precentral gyrus | R |
| 330 | 5.51 | 5.37 | 40 | 22 | 26 | middle frontal gyrus | R |
| 123 | 5.40 | 5.27 | -10 | -78 | -16 | cerebellum | L |
| 338 | 5.32 | 5.19 | 10 | -20 | 30 | callosal body | R |

| **Table 6. Interaction HMS x Anxiety (F-contrasts) (FEWc:111, c:30)** | | | | | | | |
| --- | --- | --- | --- | --- | --- | --- | --- |
| equivK | F | equivZ | x | y | z | Label | Hemisphere |
| 260 | 45.96 | 6.42 | -32 | 0 | -16 | left amygdala | L |
| 338 | 53.76 | 6.92 | -28 | -26 | -10 | left hippocampus | L |
| 223 | 54.05 | 6.94 | 14 | 42 | -8 | paracingulate gyrus | R |
| 262 | 29.32 | 5.15 | 36 | 24 | -2 | Anterior insular cortex | R |
| 146 | 65.75 | 7.60 | -26 | 32 | -2 | frontal orbital cortex | L |
| 2689 | 60.96 | 7.34 | -36 | -80 | 28 | lateral occipital cortex superior division | L |
| 3855 | 59.21 | 7.24 | 26 | 24 | 36 | middle frontal gyrus | R |
| 1337 | 52.97 | 6.87 | -28 | 4 | 26 | left caudate | L |
| 258 | 49.52 | 6.66 | 12 | -84 | -4 | lingual gyrus | R |
| 309 | 46.32 | 6.45 | 8 | -34 | -2 | brain stem | R |
| 242 | 45.20 | 6.37 | -24 | -70 | -10 | occipital fusiform gyrus | L |
| 248 | 45.10 | 6.37 | 2 | 46 | 8 | paracingulate gyrus | R |
| 123 | 42.71 | 6.20 | -10 | -18 | -6 |  | L |
| 111 | 41.60 | 6.12 | -18 | -36 | -2 | left thalamus | L |
| 1218 | 39.97 | 6.00 | 24 | -62 | -6 | lingual gyrus | R |
| 279 | 39.82 | 5.99 | 66 | -36 | 12 | supramarginal gyrus, posterior division | R |
| 137 | 39.32 | 5.96 | -40 | -50 | -36 | cerebellum left crus i | L |
| 189 | 36.90 | 5.77 | -54 | -16 | 4 | heschl’s gyrus | L |
| 284 | 36.58 | 5.75 | -20 | 18 | 0 | left putamen | L |
| 188 | 34.98 | 5.63 | -32 | -44 | -14 | temporal fusiform cortex posterior division | L |
| 118 | 34.67 | 5.60 | -44 | -28 | -8 | middle temporal gyrus | L |
| 166 | 32.47 | 5.42 | 44 | -22 | -18 | inferior temporal gyrus, posterior division | R |
| 131 | 32.29 | 5.41 | -54 | 18 | 10 | inferior frontal gyrus pars opercularis | L |
| 223 | 32.11 | 5.39 | -6 | -2 | 38 | cingulate gyrus anterior division | L |
| 153 | 31.76 | 5.36 | -48 | -14 | 22 | central opercular cortex | L |
| 250 | 31.33 | 5.33 | 22 | 2 | 10 | right putamen | R |
| 144 | 30.37 | 5.24 | 26 | -84 | -44 | cerebellum right crus ii | R |
| 129 | 29.70 | 5.19 | -44 | -72 | -2 | lateral occipital cortex inferior division | L |
| 302 | 27.95 | 5.03 | 58 | 0 | 40 | precentral gyrus | R |
| 121 | 25.52 | 4.81 | 30 | -28 | 24 | superior longitudinal fascicle | R |

| **Table 7. Interaction HMS x Anxiety (Post-hoc T-contrasts) hms(anx> non-anx) (FWEc: 130, c:18)** | | | | | | | |
| --- | --- | --- | --- | --- | --- | --- | --- |
| equivK | T | equivZ | x | y | z | Label | Hemisphere |
| 2170 | 8.64 | Inf | -32 | -2 | -16 | left amygdala | L |
| 722 | 8.03 | 7.62 | 14 | 36 | -8 | paracingulate gyrus | R |
| 247 | 6.32 | 6.11 | 36 | -2 | -12 | Mid insular cortex | R |
| 516 | 8.34 | Inf | -34 | -76 | 30 | lateral occipital cortex, superior division | L |
| 756 | 7.41 | 7.09 | 40 | -70 | 22 | lateral occipital cortex, superior division | R |
| 1246 | 7.36 | 7.05 | -24 | -66 | 4 | intracalcarine cortex | L |
| 130 | 7.15 | 6.86 | 46 | -48 | -40 | cerebellum right crus i | R |
| 601 | 6.68 | 6.44 | 44 | -30 | 6 | superior temporal gyrus posterior division | R |
| 1272 | 6.67 | 6.43 | 14 | -44 | 62 | postcentral gyrus | R |
| 385 | 6.54 | 6.31 | -58 | -18 | -12 | middle temporal gyrus, posterior division | L |
| 265 | 6.25 | 6.05 | 22 | -84 | 24 | lateral occipital cortex, superior division | R |
| 219 | 6.22 | 6.02 | -40 | -56 | -28 | cerebellum left crus i | L |
| 143 | 5.99 | 5.82 | 22 | -10 | 40 | cerebral white matter | R |
| 693 | 5.89 | 5.72 | 40 | 24 | 26 | middle frontal gyrus | R |
| 498 | 5.73 | 5.58 | 24 | 48 | 24 | frontal pole | R |
| 272 | 5.49 | 5.35 | 22 | -72 | -36 | cerebellum right crus i | R |
| 220 | 5.36 | 5.23 | -26 | -64 | -40 | cerebellum left crus ii | L |
| 142 | 4.52 | 4.44 | 60 | -14 | -14 | middle temporal gyrus, posterior division | R |

| **Table 8. Interaction HMS x Anxiety (Post-hoc T-contrasts) anx(hms> non-hms) (FWEc: 130, c:13)** | | | | | | | |
| --- | --- | --- | --- | --- | --- | --- | --- |
| equivK | T | equivZ | x | y | z | Label | Hemisphere |
| 176 | 7.53 | 7.19 | 12 | 48 | -2 | paracingulate gyrus | R |
| 205 | 7.47 | 7.15 | -38 | -50 | -38 | cerebellum left crus i | L |
| 244 | 7.34 | 7.03 | -44 | -2 | -26 | temporal pole | L |
| 3580 | 7.31 | 7.00 | -30 | 2 | 28 | left caudate | L |
| 3195 | 7.13 | 6.84 | 22 | -20 | 42 | middle frontal gyrus | R |
| 551 | 6.73 | 6.48 | 42 | -26 | 42 | postcentral gyrus | R |
| 239 | 6.63 | 6.39 | -18 | -80 | 24 | lateral occipital cortex, superior division | L |
| 205 | 6.48 | 6.26 | -24 | 54 | 28 | frontal pole | L |
| 255 | 6.44 | 6.22 | 36 | -68 | 20 | lateral occipital cortex, superior division | R |
| 130 | 6.41 | 6.19 | 66 | -34 | 12 | superior temporal gyrus posterior division | R |
| 132 | 6.09 | 5.91 | 10 | -18 | -4 | right thalamus | R |
| 282 | 6.04 | 5.85 | 36 | -12 | -24 | parahippocampal gyrus, anterior division | R |
| 135 | 4.89 | 4.79 | -16 | 12 | 58 | superior frontal gyrus | L |

| **Table 9. Interaction Beighton score x anxiety (FEWC:134, c:9)** | | | | | | | |
| --- | --- | --- | --- | --- | --- | --- | --- |
| equivK | F | equivZ | x | y | z | Label | Hemisphere |
| 846 | 9.22 | Inf | -32 | -48 | -6 | temporal occipital fusiform cortex | L |
| 144 | 6.85 | 6.59 | 4 | -62 | -40 | cerebellum vermis viiib | R |
| 195 | 6.76 | 6.51 | 40 | 30 | 8 | inferior frontal gyrus pars triangularis | R |
| 551 | 6.12 | 5.93 | 26 | -50 | -8 | temporal occipital fusiform cortex | R |
| 195 | 6.07 | 5.88 | 0 | -74 | 20 | supracalcarine cortex |  |
| 515 | 5.98 | 5.81 | -36 | 22 | -30 | temporal pole, also anterior insula in this cluster (-34,22,0) | L |
| 219 | 5.47 | 5.33 | 12 | 48 | 18 | paracingulate gyrus | R |
| 134 | 5.13 | 5.02 | -56 | -14 | -14 | middle temporal gyrus posterior division | L |
| 184 | 5.09 | 4.97 | -14 | -44 | -20 | cerebellum left v | L |

| **Table 10. Interaction Beck x HMS (FEWc: 150, c:7)** | | | | | | | |
| --- | --- | --- | --- | --- | --- | --- | --- |
| equivK | F | equivZ | x | y | z | Label | Hemisphere |
| 150 | 5.93 | 5.76 | -22 | 0 | 2 | left putamen | L |
| 36033 | 11.32 | Inf | -16 | -36 | 16 |  | L |
| 416 | 8.45 | Inf | 14 | -8 | 68 | superior frontal gyrus | R |
| 176 | 7.72 | 7.35 | 46 | -70 | -32 | cerebellum right crus i | R |
| 875 | 7.08 | 6.79 | -50 | -14 | -22 | inferior temporal gyrus posterior division | L |
| 268 | 6.14 | 5.95 | -8 | -10 | 44 | cingulate gyrus, anterior division | L |
| 154 | 5.12 | 5.00 | 22 | -38 | 58 | postcentral gyrus | R |

| **Table 11. PPI-insula: Interaction between HMS and anxiety (FEWc:174, c:2)** | | | | | | | |
| --- | --- | --- | --- | --- | --- | --- | --- |
| equivK | F | equivZ | x | y | z | Label | Hemisphere |
| 185 | 36.46 | 5.16 | 32 | -68 | 42 | lateral occipital cortex, superior division | R |
| 174 | 22.52 | 4.18 | 42 | -46 | 44 | supramarginal gyrus | R |

| **Table 12. PPI-amygdala (FEWc:207, c:1) Post-hoc T-contrasts (hms>non-hms)** | | | | | | | |
| --- | --- | --- | --- | --- | --- | --- | --- |
| equivK | T | equivZ | x | y | z | Label | Hemisphere |
| 207 | 4.17 | 3.88 | -10 | -76 | 38 | precuneus | L |
